# Communication and teamwork between dental surgeons and dental technologists during fabrication of removable complete dentures in Makerere University Dental Hospital: a qualitative research study

**DOI:** 10.1101/2025.01.03.25319953

**Authors:** David Nono, Godfrey Bagenda, Isaac Okullo, Charles Mugisha Rwenyonyi

**Affiliations:** Department of Clinical Research, Central University of Nicaragua, Managua, Nicaragua; Department of Clinical Research, Texila American University, Georgetown, Guyana; Department of Dental Technology, Makerere University, Kampala, Uganda; Department of Research, Uganda Institute of Allied Health and Management Sciences-Mulago, Kampala, Uganda; Department of Anatomy, Makerere University, Kampala, Uganda

## Abstract

**Background and Objective:** Edentulous patients seek services from dental professionals to replace lost teeth with removable complete dentures (RCDs). The dentist (dental surgeon) and dental technologists are expected to work as a team to fulfill all the expectations of the patient. Effective communication between the dentist and the technologist is important in ensuring that dentures are well-fabricated and fitted, and it also impacts the working relationship between the two professionals. The present study explored communication and teamwork between dental surgeons and dental technologists during the fabrication of RCDs.

**Design:** The study employed a qualitative design to explore communication and teamwork between dental surgeons and dental technologists. It was conducted using in-depth interviews of dental surgeons and technologists guided by data saturation.

**Setting:** The study was conducted in Makerere University Dental Hospital in Kampala, Uganda.

**Participants:** Twenty five participants: 13 dental surgeons and 12 dental technologists were interviewed during data collection.

**Results:** Generally, participants reported having a good working relationship, where they closely worked together and supported each other to ensure that patients are satisfied with RCD services. The dental surgeons and technologists used different forms of communication. However, the common form of communication was through the use of dental laboratory request forms on which all details about the procedures and materials were provided by the dental surgeons to dental technologists. To complement the dental laboratory request forms, the use of phone calls and electronic media like WhatsApp messages were equally important in emphasizing instructions and follow-ups to ensure that the work was well executed.

**Conclusion:** Most of the participants reported having a good working relationship aimed at offering the best services to the patients. Communication between the dental surgeons and dental technologists was mainly through the use of dental laboratory request forms. Phone calls and electronic media were equally important in emphasizing instructions and follow-ups to ensure that the work was well executed.

**Strengths and Limitations of this study:**

◆ The strength of the present study was that it provided baseline data on communication and teamwork between dental surgeons and dental technologists, and how best they can be improved.
◆ The study limitations were that the participants were limited to general dental practitioners and no dental consultants were interviewed who probably could have different views. It was also conducted at one site (Makerere University Dental Hospital), which renders the findings to be generalized with caution.

## INTRODUCTION

Edentulous patients look to dental professionals to replace their lost teeth^1^ to reinstate function and beauty in addition to preserving the remaining teeth^2, 3^. Removable complete dentures (RCDs) are commonly used to rehabilitate edentulous patients^4^. Patients explain to the dentist what they need, and then the dentist and dental technologist form a team to fulfill all the expectations of the patient treatment^1^. The role of the dentist is to develop a treatment plan (agreeable to the patient) and send instructions to the dental technologist^5^.

During the treatment process, effective communication between the dentist and the technologist is important^6, 7^, which leads to a well-fabricated and durable denture^8, 9^ to the satisfaction of the dentist and patient. Consequently, this results in a good working relationship between the dentist and technologist^1, 8^. Appropriate communication is important since in most cases, the dental technologists never interact with the patient^10^. In effect, dentists delegate laboratory-related procedures to the technologists based on the patient’s functional and aesthetical needs^11^. During the fabrication of dentures, the most effective communication between the two dental professionals is mainly through the use of laboratory request forms^9, 11^. Therefore, the dentist must provide clear written detailed instructions and also deliver accurate impressions to the technologists^12, 13^.

However, on the other hand, communication should not be one-way, the technologists should also share information with the dentist^14^. The technologists ought to critically observe the instructions on the laboratory request forms if the communication is clear enough to allow them to proceed with denture fabrication^11^. In cases where essential information in the laboratory request form is deficient, the technologist ought to contact the dentist to obtain more clarity^15^. Otherwise, if clarity is not sought, it results in the fabrication of an inaccurate denture, leading to increased chair side time, additional costs, and frustration for the dentist and patient^16^.

In many instances, dentists receive dentures from the laboratory that do not meet the expectations of the dentist or patient due to poor communication^17, 18^. Some studies^15, 19^ have shown that a considerable number of written instructions for denture fabrication are not properly prescribed and only a few technologists contact dentists for clarity relating to the design of the dentures. In Uganda, it is not clear how dentists and technologists communicate during denture fabrication, the challenges they face, how they affect the treatment process for patients, and how they address them. Therefore, the present study explored communication and teamwork between dental surgeons and dental technologists when fabricating removable complete dentures.

## Materials and Methods Study Design

The study employed a qualitative design to explore the experiences of communication and teamwork during RCD fabrication services among dental surgeons and technologists. The data were collected using in-depth interviews.

## Study Site

The study was conducted at Makerere University Dental Hospital in Kampala. Kampala is the capital city of Uganda. The hospital is a teaching and health service delivery facility of Makerere University. It is the largest and adequately equipped dental facility employing the highest number of dental specialists in Uganda. It has a well-established prosthetic dental laboratory and offers specialized dental services including rehabilitation of edentulous patients with RCDs, mostly to staff and students of the University, and other neighboring communities. The hospital attends to approximately 660 outpatients per month of which about 20 are rehabilitated using RCDs (Registry of Dental Records, 2022).

## Selection of Study Participants

The selected participants were registered and practicing dentistry including RCD fabrication. They were purposively selected in consideration of areas of their clinical/laboratory services. Their selection also considered variation in the duration of practice, level of training, roles in denture fabrication procedures, and fitting to ensure a fair representation of the study population. The last respondents (13^th^ dental surgeon and 12^th^ dental technologist) were determined based on data saturation, where continuing to collect more data would not yield any new information.

## Inclusion criteria

Dental surgeons and dental technologists participating in the provision of RCDs in Makerere University Hospital.

## Exclusion criteria

Dental surgeons and dental technologists who were either sick or did not consent to participate in the study.

## Data Collection

Before participating in the study, written informed consent was provided by the participants. They were assured of confidentiality such that no identifiers like names were used in data collection and preparation of reports. The research assistants personally approached the participants and requested them to participate in the study. The interviews were conducted in a conducive environment preferred by the participants, ensuring confidentiality and privacy while sharing their insights. The interview comprised open-ended questions with probes to prompt dialogue and unmediated opinions on aspects of communication and teamwork during RCD fabrication services. Data collection and the subsequent analysis were conducted as an interactive process. The last participant for IDI was established by informational redundancy, i.e., when the discussion or interview generated no new information^20, 21^. The interview for each respondent took 30 to 45 minutes and was audio-recorded in addition to note taking. This was done with the help of a trained research assistant with a background in social sciences and experience in qualitative research. The recruitment of participants started on 15^th^ September 2023 and ended on 20^th^ November 2023.

## Quality control

The data collection tool was pretested by the principal investigator and amendments were made to improve their validity and reliability. The research assistant was trained in data collection. The in-depth interviews were audio-recorded to capture any discussion that was missed in taking notes. Additional notes capturing body language and gestures during the interviews were also recorded. To guarantee reliability, four standards were applied: credibility, confirmability, dependability, and transferability. Peer debriefing and enlisting the assistance of more seasoned qualitative researchers who evaluated and provided feedback on the study technique and findings to guarantee that the data were correct and pertinent helped to establish credibility. The research background, including the characteristics of the selected participants and study setting, was thoroughly explained in the methods section for the readers to evaluate whether the findings can be applied to their contexts. The detailed description of the techniques and analysis are fully described to allow for possible replication of the study. To ensure that the conclusions of the study were unbiased and founded on the testimonies and statements of the participants, clearly defined themes and a coding scheme were used to generate codes and identify trends in the analysis^22^.

## Data Management and Analysis

Data management involved the transcription of interview verbatim recordings. After transcription, the 25 transcripts: 12 for the dental technologists and 13 for the dental surgeons were coded leading to the development of a code book. The code book was tested using five transcripts and imported into Nvivo 14 for systematic organization and analysis. After reading and re-reading the transcripts, emerging and recurrent themes were identified and subsequently interpreted. Data were analyzed using themes^23^. Personal experiences were captured as individual quotes.

## Ethical Considerations

Ethical approval of the protocol was obtained from the Makerere University School of Health Sciences Research and Ethics Committee (Reference Number: MAKSHSREC-2023-486) as well as the Uganda National Council for Science and Technology (Reference Number: HS3092ES). Permission to carry out the study was obtained from the administration of Makerere University Dental Hospital. Written informed consent was obtained from all the respondents before taking part in the study in accordance with the Helsinki Declaration^24^. All the data collected were kept securely in a cabinet under lock and key and only accessible to the investigator.

## RESULTS

The study involved 25 respondents with varying demographic characteristics (Table 1). All the respondents had either a Bachelor in Dental Surgery or a Bachelor of Dental Technology: Thirteen were dental surgeons and 12 were dental technologists. Fourteen participants were aged 26-35 years (Table 1).

**Table 1.**
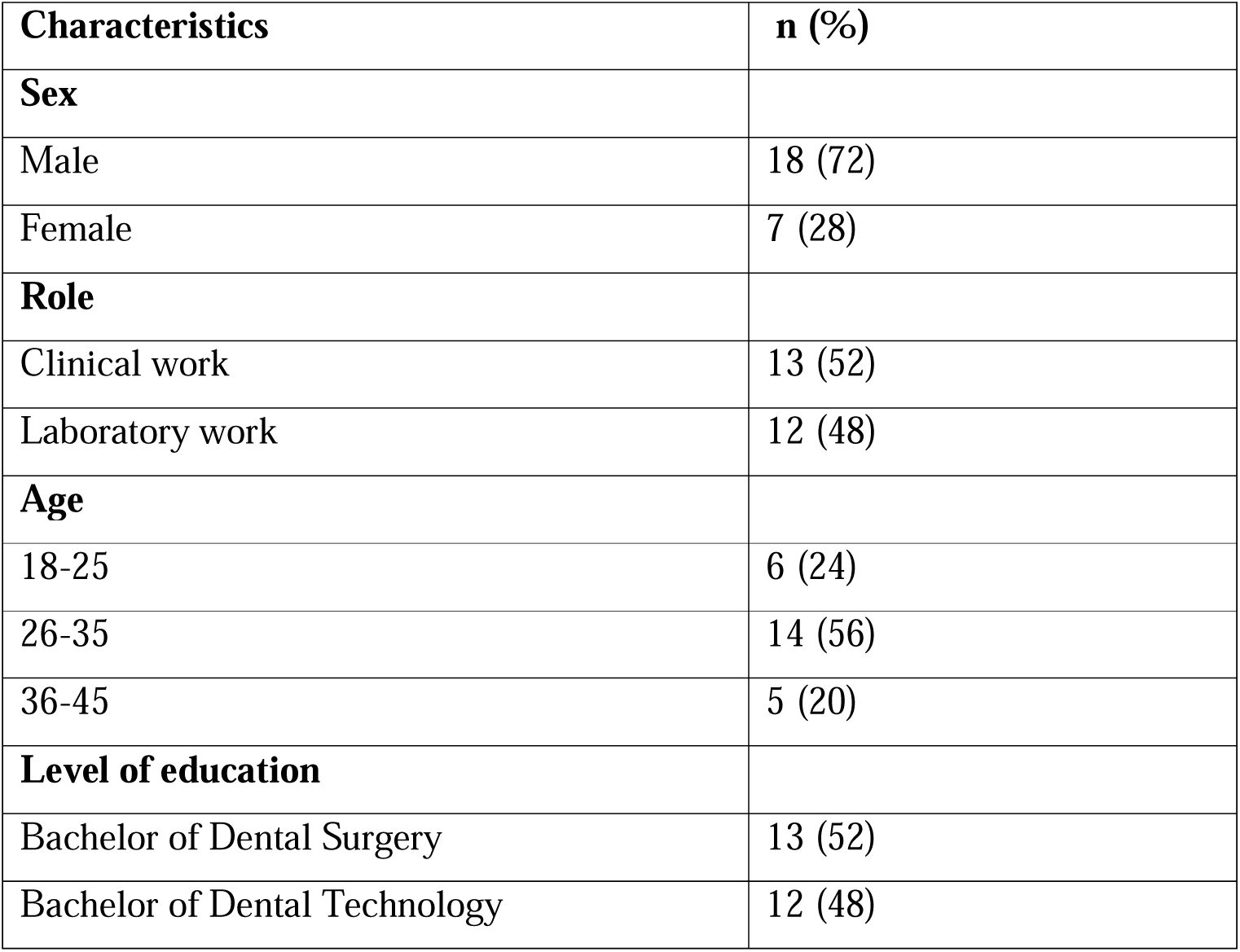
The frequency distribution of the respondents according to their social demographic characteristics (N=25)

| **Characteristics** | **n (%)** |
| --- | --- |
| **Sex** |  |
| Male | 18 (72) |
| Female | 7 (28) |
| **Role** |  |
| Clinical work | 13 (52) |
| Laboratory work | 12 (48) |
| **Age** |  |
| 18-25 | 6 (24) |
| 26-35 | 14 (56) |
| 36-45 | 5 (20) |
| **Level of education** |  |
| Bachelor of Dental Surgery | 13 (52) |
| Bachelor of Dental Technology | 12 (48) |

Four themes emerged from the data (Table 2).

**Table 2:** Themes and subthemes.

| **Theme** | **Subtheme** |
| --- | --- |
| Communication and teamwork experiences of dental practitioners | • The importance of communication in the management of edentulous patients<br>• Communication and teamwork experiences |
| Modes of Communication | • Written Laboratory request forms<br>• Phone Call reminders<br>• Digital information-sharing platforms |
| Communication challenges during complete denture fabrication | • Unclear and incomplete information requests<br>• Delayed laboratory request forms<br>• Not following instructions |
| Ways to improve communication and work relationships | • Active listening<br>• Encourage more of written information<br>• Ensure lab request forms are available<br>• Follow-up on requests made and seek clarity<br>• Use of multiple means of communication |

## Communication and teamwork experiences of dental practitioners

### The importance of communication in the management of edentulous patients

Participants contended that communication is a critical component of complete denture fabrication procedures. They reported that communicating effectively what you expect from a dental technologist through writing, is very important as revealed in the narrative below:

*“Communication is very important, writing down the details [of what] you want is very important, what you expect from a technologist because s/he needs to know what to expect of him or her. So, communication is very paramount” (P004_dental surgeon)*

Another participant emphasized that, if communication is effective, it will help prevent any likely mistakes that can be made during denture fabrication procedures.

*“Communication is key because if you miss out on anything in writing or discussion, it will affect the outcome of denture fabrication” (P019_Dental surgeon)*.

### Communication and teamwork experiences

We asked our participants to narrate their communication and teamwork experiences while working with colleagues when managing patients. The majority reported very positive working relationships with a few who reported some challenges. Those who reported positive experiences were inclined to the fact that they sit and discuss issues amongst themselves and help each other to find solutions.

*“Of course, I work together with a dental technologist and at times we discuss because he has his knowledge and I have mine. In the discussion, we help each other. At times we discuss to see what is ideal and suitable for a certain patient.” (P014_Dental surgeon)*

Others offer on job support to their colleagues in cases where one does not know what to do either because of limited expertise or issues of being fresh in the field.

*“I try my best to direct them [dentists] on the procedure in case they are not so sure…I try my best in case they don’t do it well. I can only tell them ‘you did not do this well’, so then they either repeat it or they ask for help” (P006_Dental Technologist)*.

Sometimes the nature of good working relationships is expressed at times when wrong procedures are taken and realize later that they need to be repeated. The majority reported taking it in good faith to repeat the procedures because they all focus on giving their patients the best of services.

*“…if there is a problem, I sit down and find a solution with the technologist. We always run back to each other. Even if it means repeating the whole process, we always repeat it because, at the end of the day, we want the best outcome for the patient. So, no one should be the boss of the other. We must always produce good work” (P018_Dental Surgeon)*.

Working together and emphasizing good working relationships was a safety measure to ensure that issues to do with forgetfulness due to too much work are avoided because they will be reminding each other.

*“At times because of being busy, you can forget something very minor, but I believe if you work hand in hand with your technologist, anything you leave out s/he can remind you. Alternatively, if the technologist leaves out something, the dentist can correct him/her” (P018_Dental Surgeon)*.

## Modes of Communication

We explored the means of communication used by dental practitioners to communicate with each other. They all mentioned four commonly used means of communication including; written prescriptions, phone calls, face to face and sometimes digital communication as presented below.

### Written Laboratory request forms

The common and most effective means of communication used was written information on the laboratory request forms. These forms entail a detailed description of the information the dentist deems important based on the patient complaint(s) presented in the clinic, the diagnosis and procedures to be taken by the technologist to meet the expectation(s) of the patient. In other words, it is the dental surgeon that uses the laboratory request forms to write procedural instructions to the technologists.

*“One means of communication is through writing, what we call laboratory request forms sent to the laboratory, where the clinicians write all the details that they want the technologist follow to fabricate dentures” (P010_Dental Technologist)*.

*“I write instructions on a laboratory request form to the technologist.” (P004_dental surgeon)*.

Also, written prescriptions are used in situations where the needed procedures are not straightforward. Yet, in cases where the procedures to be done are obvious, phone calls are used to emphasize and simply remind the colleagues to act up on them.

*“At times we write down, but alongside the written prescription is supported with a phone call. Then yes, there are those instances when you know it is straightforward, you just call the technologist and direct what to do” (P018_Dental Surgeon)*.

### Phone Call reminders

Once the instructions are delivered, s/he leaves behind a phone number through which s/he can be reached to discuss any other needed information. Also, in cases where a person thinks that what they have prescribed might be ignored by their colleagues, they make follow-up calls to make clarifications.

*“Of course, part of the communication most times is written, other times in case something is not clear to you, you give them a call and ask for more clarity” (P006_Dental Technologist)*.

*“I also call the technologist. I am in direct contact with the technologists at every stage of the procedure because for you to get good work that is the way to go” (P004_Dental surgeon)*.

Phone calls are also seen to be effective, especially when it comes to discussing other issues related to patients’ expectations and information that may not be documented.

*“We always use phone calls and then a few writings. We usually use phone calls when we are trying to give the history and expectations of the patient, and all that” (P023_Dental surgeon)*.

One participant admitted that majority of dentists use phone calls and a few write down the information.

*“A bigger percentage of dentists use phone calls. A smaller percentage take time to write down their instructions, specifically, what they expect you to do and what they want” (P012_Dental Technologist)*.

### Digital information-sharing platforms

Some respondents reported that they use digital platforms to communicate with each other. Especially if the instructions are hard to understand. One can take a video while demonstrating what exactly should be done and share it with a colleague to follow and do the right procedures.

*“Normally, I explain to them what I need. I explain those procedures how they should be done. I take videos and send to them. May be if the problem was impression taking, I can record a video of myself taking a good impression” (P008_Dental Technologist)*

*“…. then some [dental surgeons] send photos to accompany what they have written down” (P010_Dental Technologist)*

## Communication challenges during complete denture fabrication

Despite the very emphasis enshrined in the importance of communication and the available effective means of communication, a number of challenges still prevail and if not addressed,

can hinder effective and successful complete denture fabrication. There are four major challenges including; unclear and incomplete information requested, missing information, delayed lab requests, and misinterpreting instructions. These are presented below:

### Unclear and incomplete information requests

Some participants especially the dental technologists narrated how they are sometimes given unclear information about denture fabrication. They reported that the only information normally specified to them is the shade of the teeth and colour of the gum, and they are expected to make all other choices like size and length of teeth on their own, which to the technologists is a communication gap that needs to be addressed.

*“Of course, the communication is not elaborative. They [clinicians] just give me the shade of teeth and colour of gum, and that’s it. They expect me to do other things by myself, they don’t specify the mould or the tooth material, tooth size and the length of the teeth.” (P008_Dental Technologist)*

As a result of incomplete information provided, the technologists find it hard to determine the actual measurements to follow and those who try to reach out to their colleagues, they cannot give them accurate information because of the recall bias, and yet, at the same time, the patient has gone back home. This makes the whole process complicated for the practitioners.

*“On many occasions it is us [technologists] to ask them [clinicians], when they have sent work, but they do not write anything. Most of them [clinicians] don’t write so we have to call them and of course, as we are calling we may not get the exact details. Remember by the time we are calling to receive the measurements, the patient will have already gone” (P006_Dental Technologist)*.

### Delayed laboratory request forms

Some technologists reported to have been frustrated by some clinicians who delay to submit the laboratory requests, and this means that the technologists will have nowhere to base to fabricate the dentures.

*“It would be fair to say some clinicians have really failed to write laboratory request forms. They’re supposed to give us forms detailing instructions, but they have failed*.

*You have to call [and ask], which tooth shade, gum colour etc?” (P011_Dental Technologist)*

### Not following instructions

The frustrations with communication were shared amongst all the categories of professionals. Similar to the technologists, the surgeons too complained about the technologists not following instructions and opting for easy ways or shortcuts to the procedures of denture fabrication. The challenges which they described as a liability.

*“So, some of the technologists like shortcutting, which is a big liability, and some of them, you ask for one thing, they do the other not following the instructions. They don’t follow the instructions or the prescription from the dentist. Sometimes they skip some steps” (P021_Dental Surgeon)*

## Ways to improve communication and work relationships

### Active listening

The participants emphasized a number of strategies they thought would help improve communication and teamwork. One of these is the ability to listen to each other which will translate into good service delivery.

*“The team [dentist and technologist] must be compatible with each other, it is easier to communicate when you listen to each other‘s viewpoints. When one listens and one communicates then you let the other party do the good thing.” (P002_Dental surgeon)*.

### Encourage more of written information

Because of challenges related to recall bias and the gap in the use of phone call reminders, it was suggested that the best way to improve communication is to emphasize written information that a person can refer to any time they want to conduct a procedure.

*“I believe the best way to communicate is by writing. Writing is the best way because there, it’s like an agreement. But if it’s by mouth, someone can forget. They can forget so I think the gold standard would be writing, but also supplemented communicating through phone calls” (P018_Dental Surgeon)*.

Some participants discouraged sole communication via phone calls, mentioning that it’s unreliable means of communication, linking it again to recall bias. They emphasized that the chances of having the right procedures is dependent on one’s ability to remember what s/he was told on phone.

*“Sole communication on phone is a very bad method to rely on, it is better to have something written and then you emphasize it on the phone call. You may call me on the phone and you tell me ABCD, I tell you it’s okay, but by the time I come back to do the thing I have forgotten what you have told me.” (P022_Dental Technologist)*

### Ensure lab request forms are available

As a way of harnessing the use of written information, participants advised that there is a need to design the laboratory request forms and have them availed to the staff for use in case of need. If these forms are availed, it becomes easier to make the necessary follow-ups in case of omissions or any errors.

*“I make sure that Laboratory Request Forms are available in the clinic for clinicians to fill when they are with patients, not in the lab, because when it’s in the lab, they [clinicians] will [say] they don’t have time to come up there. So, I put them in the clinic. And I do not accept work without making sure that the dentist has written a request form because I need to have records on my side” (P011_Dental Technologist)*.

### Follow-up on requests made and seek clarity

One way to improve communication was to ensure that the instructions given on either laboratory request forms or phone calls or even face to face are clear to the rest of the staff. And in cases where the information is not clear, there should be room to adjust and seek for clarity.

*“When they [clinicians] send me work without clear instructions, I always try and call them back. I advise them to always write down instructions and ask for any information that I don’t understand about the case they have sent me” (P012_Dental Technologist)*.

In this regard, there is a need for continued and constant communication with colleagues as it helps to cement the teamwork, to learn from each other and improve the practices, and service delivery.

*“You have to frequently talk to colleagues to share knowledge. I don’t think one would work alone. Well, it is possible, but of course there is something you learn from your colleagues when you share experiences with them and work with them” (P017_Dental Surgeon)*.

### Use of multiple means of communication

Because of the reported challenges with the use of particular means of communication, it was advised that there should be the adoption of multiple means of communication. This will help in a way that if one means of communication is not effective, its weaknesses can be compensated with another.

*“Minimizing the gap, normally is by written instructions. That’s number one, number two, also following up with phone calls or WhatsApp messages, then in case they have any doubt let them call. We always give that option to them (P021_Dental Surgeon)*.

## DISCUSSION

This study explored communication and teamwork between dentists and dental technologists during the fabrication of RCDs. Teamwork was important given the differences in experience, knowledge, and expertise among the dentists and technologists. Bringing together their expertise and knowledge ensured that they met the patient’s expectations. The relationship between dentists and technologists ought to be an interactive one. There has to be full participation and careful management of information for this kind of collaboration to grow for the benefit of the dental professionals as well as the patients^25^.

The present study also found that working together provided some form of checks and balances. When dentists kept in touch with technologists ensured that dentures were made according to the plan as per the prescriptions of the work. Similarly, previous findings show that alignment of expectations and means of communication employed are key in shaping the dentist-technologist work relationship^26^. A strong dentist-technologist relationship is built on having mutually shared goals and both of them should have the desire for better results and willingness to make it happen^27^. Otherwise, a weak collaboration between dental professionals can result in the production of poor prostheses and eventually substandard patient care services^26^.

The present study reported use of laboratory request forms as the common means of communication, which were used by dentists to explain to technologists the procedures and details of the work to be done. One of the advantages of using laboratory request forms is that the details are written and one can always refer to them whenever needed. However, the challenge may be when the instructions are confusing, unclear, or misinterpreted by the technologist. In the present study, deviation from the instructions to the technologists was pointed out by the dentists. Other studies^28^ have identified challenges of using paper-based forms of communication. Written instructions reportedly lead to miscommunication between the technologist and dentist.

However, in other cases, a dentist may not adequately communicate the instructions to the technologist. Although the laboratory request forms are expected to be detailed, the technologists reported that the information included on the laboratory request forms was insufficient for instance, limited to tooth shade and gum color, and this would result in making inadequate dentures. Similarly, Bashir et al.^29^ found that dentists many times have to re-order the prostheses because of errors made in the laboratory, even when the prescriptions are clear due to deviation from the desired materials and procedures resulting from misinterpretation of the information on the laboratory request form. Relatedly, another study^27^ found that dentists many times write a few notes about the patient and when technologists rely on such information to make a restoration, it ends up not being satisfactory to the patient.

Similarly, other studies^11, 30^ found that work authorizations sent by dentists were inadequate or incomplete. This was also the case in a study by Ismail & Al-Moghrabi^26^ where technologists reported that insufficient information in the laboratory request forms led to unexpected delays, repetitions, or repairs resulting in loss of revenue and clinical time and compromised quality of patient care. Poor interpretation of the written instructions implies that there could be a problem with terminologies or insufficient details which makes the technologist to guess. Thus, the dentist should write as clearly and elaborately as possible to ensure that the technologist does what is expected of him or her. Additionally, when instructions are clear, it minimizes delays and enhances accuracy.

In the present study, respondents emphasized open communication. Communication should not only be one, but two-way; technologists should reach out to dentists if something in the laboratory request form seems unclear. Schoenbaum & Chang^27^ revealed that many times the level of communication between dentists and technologists can be a weak link in the process of making dentures. Technologists should be able to inform dentists if they have any issues relating to the preparation of teeth, the design of dentures, or the quality of the impression^31^.

A two-way communication leads to better information, adequate preparations, and improved service delivery^27^.

The respondents in this study emphasized that using phone calls was necessary to remind their colleagues or to emphasize some of the instructions in the laboratory request forms. It also ensures that dental professionals are on the same page in terms of what needs to be done. On the contrary, other studies^15, 19^ found that only a few technologists contacted dentists for clarity relating to the design of the dentures.

In the present study, respondents revealed that phone calls were also important for technologists to ask for any additional information in case they felt some information was not included on the laboratory request forms. These follow-up phone calls were also important in maintaining and strengthening teamwork among the dentists and technologists. In case they felt they needed to demonstrate specific activities, digital platforms such as WhatsApp messages were effective since one could make a demonstration and share a video, especially for complex procedures. Similarly, in one study^26^, technologists highly rated the use of visual aids, and verbal discussions either face-to-face or virtual audio-visual in addition to written prescriptions. Similar to written documentation, video scripts are important for reference

purposes.

The advantage of phone calls over written information is that through a phone call, one can discuss at length and ask for more clarity or discuss something about the patient so that they understand better the patient’s condition or characteristics so that they design a prosthesis that will satisfy the patient. Communication using all these modes was key in ensuring that the right information was shared among the dental professionals so that they were aware of what they were supposed or expected to do. Better communication approaches help to minimize errors, save time, and improve the quality of the final denture^28^. On the other hand, the inability to seek clarity would imply a lack of teamwork.

Given the challenges expressed in the present study, respondents felt using all methods at their disposal would improve communication and complement each other. In case less information was shared through one method, another one would enable providing additional and clearer information. For this to be possible, flexibility is important so that dental professionals are not restricted to one method, although written instruction is the dominant method. The ability to use different means of communication requires additional resources for effective implementation.

## Conclusion

Most of the respondents reported having a good working relationship and they always engaged and supported each other to offer the patients the best services. When it came to making restorations for the patients, communication between the dentists and technologists was mainly through the use of laboratory request forms on which all details of the prostheses were written. However, the use of phone calls and electronic media was equally important in emphasizing what had to be done and following up to ensure that the work was well executed.

## Implications for practice

The creation of a conducive environment that supports open and free communication is important as it supports teamwork and allows dental professionals to exchange information about the edentulous patient’s condition and what needs to be done for adequate rehabilitation. In case anything is not clear to the technologists, they must refer back to the dentists so that whatever they do not understand is explained to them. There is a need to improve on the information written in the dental laboratory request form. For instance, there is a need for consensus on what important information should be included in the template dental laboratory request form to ensure effective service delivery.

## Recommendation

Clear and well written laboratory request forms indicating what should be done are encouraged to streamline service delivery. This should be supplemented with other channels of communication like phone calls and social media platforms.

## List of Abbreviations

RCDs - Removable complete dentures

## Declarations

### Ethics approval and consent to participate

Ethical approval of the protocol was obtained from the Makerere University School of Health Sciences Research Ethics Committee (Reference Number: MAKSHSREC-2023-486) as well as the Uganda National Council for Science and Technology (Reference Number: HS3092ES). Permission to carry out the study was obtained from the administration of Makerere University Dental Hospital. Written informed consent was obtained from all the participants who took part in the study. The purpose of the study was explained to the participants and their participation was voluntary. Their agreement to participate in the study did not waive their rights in any way and this was in accordance with the Helsinki Declaration^24^. All the data collected were kept securely in a cabinet under lock and key and only accessible to the investigator^22^

## Patient consent for publication

Not applicable

## Data availability statement

No data are available.

## Patient and Public involvement

Patients’ and the public were involved in the design, conduct, reporting, and dissemination plans of this research. More details can be got from the Methods section.

## Provenance and peer review

Not commissioned; externally peer reviewed.

## Author’s contributions

DN, IO, MA, GB, and CMR participated in the conception of the study, study design, data analysis and manuscript preparation. DN and GB participated in data collection. All authors drafted the manuscript and approved the final version.

## Data Availability

No data are available. All data produced in the present work are contained in the manuscript

## Acknowledgments

The authors are grateful to the Government of Uganda through the Makerere University Research and Innovations Fund Secretariat for supporting this study and participants for their willingness to participate in the study.

## Open access

This is an open access article distributed in accordance with the Creative Commons Attribution Non Commercial (CC BY-NC 4.0) license, which permits others to distribute, remix, adapt, build upon this work non-commercially, and license their derivative works on different terms, provided the original work is properly cited, appropriate credit is given, any changes made indicated, and the use is non-commercial. See: http://creativecommons.org/licenses/by-nc/4.0/.

## ORCID iDs

David Nono http://orcid.org/0009-0007-8443-7582

Charles Mugisha Rwenyonyi http://orcid.org/0009-0002-5936-8195 Isaac Okullo http://orcid.org/0009-0002-3766-5244

Godfrey Bagenda http://orcid.org/0009-0008-8860-3194

